# Prospective multicenter validation of a new laboratory workflow integrating the free light chains kappa quotient in CSF analysis-Protocol of the ORCAS study

**DOI:** 10.1101/2023.07.16.23292727

**Authors:** Alexander Dressel, Malte Johannes Hannich, Marie Süße

## Abstract

**Introduction:** Free light chain kappa (FLCκ) are a newer sensitive biomarker to detect intrathecal immunoglobulin synthesis. Here we report the study protocol of the ORCAS study which will evaluate a novel laboratory work flow recently proposed including FLCκ analysis simplifying cerebrospinal fluid (CSF) analysis.

**Methods:** The ORCAS study is a prospective multicenter diagnostic biomarker study across at least 4 study sites. The study protocol does not specify which assays should be applied in the participating laboratories to detect oligoclonal bands (OCB) or measure FLCκ or Ig synthesis to represent real world data. Primary outcome parameter is the sensitivity and negative predictive value of the absence of local FLCκ synthesis for the absence of intrathecal synthesis according to OCB and/or intrathecal IgG, IgA, IgM synthesis in quotient diagrams. The reference range of FLCκ will be indicated by the FLCκ quotient diagram. This study was designed according to the STARD criteria.

**Perspective:** The establishment of a newer biomarker in routine practice is associated with significant difficulties, such as the inconsistent comparability of different measurement platforms, the establishment of suitable cut-offs or insufficient knowledge regarding sensitivity and specificity of the biomarker. If the ORCAS study objective is achieved, the use of the proposed workflow integrating FLCκ analysis in routine practice in CSF diagnostics could help to better characterize the intraindividual and disease-specific intrathecal humoral immune response in a resource-efficient manner.

## Introduction

The presence of locally synthesized immunoglobulin (Ig) in the cerebrospinal fluid (CSF) is a biomarker for central nervous system (CNS) inflammation and a basic analysis in CSF diagnostics. In clinical routine, Ig synthesis is assessed by quantifying intrathecal IgG,< IgA and IgM using the Ig CSF/serum ratio in relation to the albumin CSF/serum ratio in a respective quotient diagram (1). However, the most sensitive biomarker for intrathecal IgG synthesis is the qualitative determination of oligoclonal bands (OCB), which utilize isoelectric focusing to identify IgG clones seen in CSF but not in serum (2). More recently a newer biomarker for intrathecal Ig synthesis, the free light chain kappa (FLCκ) have been introduced in CSF analytics (3). The analysis of FLCκ has several advantages over the previous reference standard, OCB analysis, such as faster and objective quantitative results, simpler application and cost savings (4-6). FLCκ are produced in excess during Ig synthesis of all Ig classes and released into serum and CSF (7). There have been different approaches for establishing a reference value, such as the FLCκ index (5,8-9). The index is determined by the FLCκ CSF/serum quotient in relation to the albumin quotient. The diagnostic sensitivity to detect intrathecal inflammation using this approach is comparable to OCB at least in multiple sclerosis patients (3,5,9). However, there is still disagreement on which cut-off should be used for the FLCκ index. This hampered generalizability of the results and hindered transfer into clinical routine. Reiber et al. provided a quotient diagram in analogy to the established quotient diagrams for IgG, IgA and IgM which can now be used as a definite basis to identify an intrathecal synthesis of FLCκ (4). Using this quotient diagram we have shown in a monocentric study that the absence of intrathecal FLCκ synthesis predicts the absence of intrathecal Ig synthesis as determined by IgG, IgA, IgM quotient diagrams or the presence of OCB in CSF. In contrast, not all patients with intrathecal FLCκ synthesis have positive OCB or detectable intrathecal Ig synthesis in the quotient diagrams (10). We have therefore suggested to adapt the CSF laboratory analysis using the FLCκ quotient as an initial step to identify those CSF-serum sample pairs that require additional analysis with Ig quantification and OCB analysis. Among other aspects, this approach is necessary because the complete replacement of the previous reference diagnostic, OCB and Ig synthesis, for cost and simplicity reasons by FLCκ would lead to a number of diagnostic errors.

## Methods

### Aim of the trial

The current study is designed to determine whether the absence of intrathecal FLCκ synthesis according to the quotient diagram for FLCκ predicts the absence of quantitative or qualitative Ig synthesis in CSF.

This study is necessary in order to transfer this newer biomarker into clinical routine in a resource-oriented but considerate manner to avoid drawing false conclusions from diagnostic uncertainties. Due to the multicenter and prospective approach, generalizability, representability and feasibility of the results can be expected.

### Study description and study design

This prospective multicenter diagnostic biomarker study across at least 4 study sites will include a total of at least 500 consecutive samples of lumbar CSF/serum pairs irrespective of the clinical diagnostic question. Routine diagnostics including Ig synthesis and OCB analysis will be performed in each laboratory according to local standards. The biomarker in question, FLCκ, will also be measured according to local standards. These additional data will be measured solely for the sake of the study and will not be reported to the clinician. At the end of the study the data will be reported pseudonymized to the main study site for final analysis. To evaluate the feasibility of this approach in clinical routine across laboratories the study protocol does not specify which assays should be applied in the participating laboratories to detect OCB or measure FLCκ or Ig synthesis. Table 1 lists the data to be reported.

**Table 1.** Data to be reported.

|  |
| --- |
| Cell count in CSF (/μl) |
| Lactate concentration in CSF (mmol/l) |
| Albumin quotient |
| IgG quotient |
| IgA quotient |
| IgM quotient |
| OCB method, OCB pattern (I-V), OCB bands in pattern II and III, OCB result (positive/negative) |
| FLCκ method, assay type, FLCκ serum and CSF concentration, FLCκ index, FLCκ intrathecal fraction (IF) |
| Creatinine in serum mg/dl |

CSF cerebrospinal fluid, Ig Immunoglobulin, OCB oligoclonal bands, FLCκ free light chains kappa

The reference range of FLCκ will be calculated using the established FLCκ quotient diagram (4). In these quotient diagrams, Ig and FLCκ CSF/serum quotients (QIgG, QIgA, QIgM, QFLCκ) are plotted in reference to the albumin CSF/serum quotient (QAlb). The formulas used for the calculation are analogous to Reiber et al. 2019 (4) (table 2) where the upper line (Qlim) of the reference range discriminates between the blood derived- and intrathecally synthesized-Ig and FLCκ fraction (10).

**Table 2.** Formulas used for calculating the reference range of the IgG, IgA, IgM and FLCκ quotient.

|  |
| --- |
| $Q\_lim(IgG) = 0.93 \times \sqrt{((QAlb)^2 + 6 \times 10^{-6})} - 1.7 \times 10^{-3}$ |
| $Q\_lim(IgA) = 0.77 \times \sqrt{((QAlb)^2 + 23 \times 10^{-6})} - 3.1 \times 10^{-3}$ |
| $Q\_lim(IgM) = 0.67 \times \sqrt{((QAlb)^2 + 120 \times 10^{-6})} - 7.1 \times 10^{-3}$ |
| $Q\_lim(FLC\kappa) = 3.27 \times \sqrt{((QAlb)^2 + 33 \times 10^{-6})} - 8.2 \times 10^{-3}$ |
| $Ig/FLC\kappa-IF = [1 - Qlim(Ig/FLC\kappa)/QIg/QFLC\kappa] \times 100 [\%]$ |

Lim: upper normal limit of the reference range, Ig Immunoglobulin, Alb albumin, FLCκ free light chains kappa, IF intrathecal fraction, Q quotient

The workflow to be tested is shown in figure 1. In detail this workflow includes the following steps: After determination of the routine CSF analysis (cell count, lactate, total protein) the determination of the FLCκ IF follows. If the FLCκ IF is negative, no further Ig or OCB analysis should be necessary if the negative predictive value of the FLCκ IF is sufficiently high. A prerequisite is that the FLCκ serum values are not above the upper limit of the reference value, as this can lead to false negative FLCκ values. If the FLCκ IF is positive, further analysis of IgA, M, and G including OCB analysis is necessary to identify disease-typical patterns of the intrathecal humoral immune response.

**Figure 1 (10).**
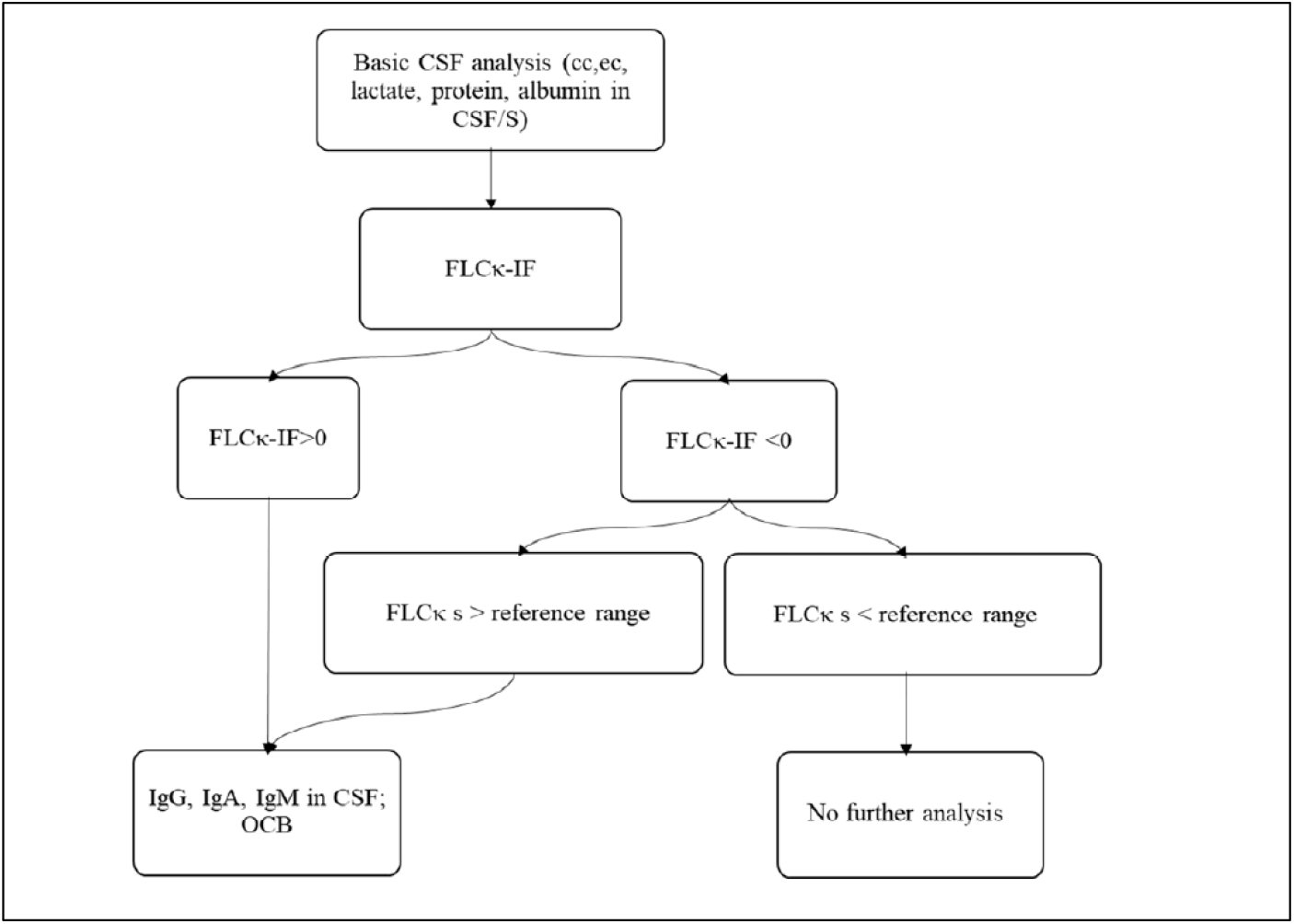
Workflow to be tested in this multicentric, prospective diagnostic biomarker study. After determination of the routine CSF analysis the determination of the FLCκ IF follows. If the FLCκ IF is negative, no further Ig or OCB analysis should be necessary provided the FLCκ serum levels are not elevated. If the FLCκ IF is positive, further analysis of IgA, M, and G including OCB analysis is necessary to identify disease-typical patterns of the intrathecal humoral immune response. CSF cerebrospinal fluid, CC cell count, EC erythrocyte count, S serum, FLCκ free light chains kappa, IF intrathecal fraction, OCB oligoclonal bands

### Arms and interventions

None.

### Outcome measures

Primary outcome parameter

The primary outcome parameter is the sensitivity and negative predictive value of the absence of local FLCκ synthesis for the absence of intrathecal synthesis according to OCB and/or intrathecal IgG, IgA,IgM synthesis in quotient diagrams.

The hypothesis to be tested is as follows: The FLCκ IF has a sensitivity and negative predictive value of at least 95% for the detection of intrathecal immunoglobulin synthesis (defined by QIgG, QIgA, QIgM > QlimIgG,A,M or detection of CSF specific OCB).

Secondary outcome parameters

1. The predictive value of the absence of local FLCκ synthesis for the absence of OCB determined by silver gel analysis compared to agarose gel
2. Predictive value of the absence of local FLCκ synthesis for the absence of OCB depending on the FLCκ assay used
3. Comparison of sensitivities to detect quantitative or qualitative Ig synthesis between index evaluation with different cut-offs (5,8-9) and the IF FLCκ
4. Analyzing material and personnel costs as well as turn-around time to evaluate the economical benefits of the proposed work flow

### Statistical analysis

A calculation of the minimum required sample size was based on the precision of two-sided confidence intervals. Since we use sensitivity and negative predictive value as co-primary endpoints, we decided to use 97.5% confidence intervals (global alpha ≤ 0.05). Precision was set at 0.05 as the maximum confidence interval width. With a sample size of 424, a two-sided 97.5% confidence interval with a width of 0.05 (0.921-0.971) is obtained in each case when sensitivity and negative predictive value of FLCκ are 95%, respectively.

The variables mentioned will be analyzed descriptively. Unless otherwise stated, all values will be given as mean values (± standard deviation). Nominal data are given as percentages. The normal distribution of the data will be tested using Kolmogorov-Smirnov analysis. Mean comparisons are made for non-parametric data using the Wilcoxon test, and for parametric data using the t-test. Comparison of nominal data is done via Fisher’s Exact Test or Chi^2^ test. A p-value of <0.05 will be considered significant. For post hoc tests, a Bonferroni correction will be applied. Sensitivity, specificity, positive and negative predictive value of the FLCκ IF are evaluated using contingency tables with 95% confidence intervals. A separate analysis is performed for the factors influencing renal function impairment and serum FLCκ concentration on the FLCκ IF. First, the number of false-negative FLCκ IF (OCB positivity or QIgG, QIgA, QIgM > Qlim) findings due to renal function impairment (GFR<89 ml/min/1.73m^2^ or creatinine values > 2 mg/dl) and/or increased serum FLCκ concentrations (> 22.4 mg/L) is described descriptively. Furthermore, logistic regression analyses are performed to investigate these factors influencing the FLCκ IF. Furthermore, a subgroup analysis is performed separately for the two OCB methods used (silver gels and immunofixation). The number of true positive (FLCκ positive, OCB and Ig positive) and false negative (FLCκ negative, OCB and Ig positive) results will be compared.

### Eligibility criteria

All consecutive paired lumbar CSF and serum samples of patients > 18 years in the respective CSF laboratories will be included. Samples with suspected artificial intrathecal Ig synthesis due to blood contamination with IF IgM > IF IgA > IF IgG and negative OCB will be excluded.

### Contacts

None.

### Perspective

Translation into routine clinical practice following the discovery and development of new biomarkers in research is often associated with significant obstacles. Difficulties are often caused by the inconsistent comparability of different measurement platforms, the establishment of suitable cut-offs and the establishment of quality control mechanisms.

The ORCAS Study will investigate the recently proposed laboratory work flow (10) that uses the intrathecal FLCκ synthesis as an initial step in order to identify samples that require additional CSF analysis. With this workflow, it is possible to characterize the intraindividual and disease-specific humoral immune response in a resource-efficient manner.

The most important aspect of this study is the prospective, multicenter and consecutive study design compared to many other FLCκ biomarker studies. Many study results are based on retrospective biobank samples with corresponding limitations regarding their application in routine clinical practice (11-14). Although most studies on FLCκ have been applied to samples from MS patients (3,6,8-9,14), FLCκ are not a specific biomarker for MS but indicate general intrathecal Ig synthesis (13). Therefore, they also have a role in a variety of other inflammatory and infectious CNS diseases (11,15-16). This aspect is taken into account in the present study, as consecutive samples will be analyzed independently of the indication for CSF analysis. Therefore, the primary outcome is also not the detection of a specific disease, but the presence of Ig synthesis according to previously established methods. When a diagnostic biomarker is available, it is important to consider specificity in addition to sensitivity. Since it is known that FLCκ can also be positive in a number of non-MS diseases, focusing on one disease can lead to diagnostic misconceptions. If FLCκ are integrated into routine diagnostics, better and more accurate characterization of the intrathecal humoral immune response in a variety of inflammatory and infectious CNS diseases may become possible.

Another important aspect of the present study will be to investigate “real word data” and thus the general applicability in routine diagnostic procedures of this biomarker. For this reason, different OCB methods and FLCκ assays will be tested depending on local standard procedures. Since this aspect is important to show how measurements are made and work at different sites, we see this variability in methodological design as an advantage.

A major problem in the routine use of new biomarkers is the choice of the correct reference range. With respect to the FLCκ, there are a large number of evaluation proposals and cut-off values (as referenced in (3)). These range from laboratory-specific to multicenter disease-specific cut-offs (5,8). However, since this in turn leads to limited comparability and diagnostic uncertainty, it would be important to agree on a reference range. Here, the hyperbolic reference range according to Hansotto Reiber (4) is the most correct from a pathophysiological point of view. This application has 2 key advantages: First, it generates a reference range with a well-defined and validated cut-off. Secondly, it can be easily applied to all infectious and inflammatory CNS diseases. The latter has the advantage of not generating false positive or false negative results with too low or too high QAlb values. All these aspects are tested in the present study, in which different evaluation methods (f.e. FLCκ index with different cut-off values) are compared with the IF of FLCκ based on the FLCκ quotient diagram.

The proposed laboratory pathway uses the FLCκ IF as the primary marker to select for further Ig analysis (IgG, A, M and OCB). There are three important limitations to this approach:

As the sensitivity of FLCκ is reduced in excessively increased serum FLCκ concentrations, samples with FLCκ serum above the upper level of normal will be excluded. This means, that in these cases (i.e. FLCκ serum above the reference range) all subsequent Ig analyses are neverthertheless necessary. Secondly, blood admixture to CSF is known to result in false positive intrathecal Ig quotient diagrams with intrathecal synthesis of IgM>IgA >IgG. We have previously shown that intrathecal blood admixture does not result in artificial intrathecal FLCκ synthesis (11). Therefore, samples with blood admixture and intrathecal synthesis of IgM>IgA >IgG that lack intrathecal FLCκ synthesis should not be considered false negatives. At last, when using the workflow, the information about the incidental presence of a monoclonal pattern in isoelectric focusing (i.e. pattern V) is lost. Since this pattern is very rare and a targeted diagnosis with e.g. immunofixation in serum is more advantageous than incidental findings, this point is negligible from our point of view (12).

### Conclusion

The study is intended to contribute to the useful and knowledge-based application of FLCκ in routine analysis in CSF laboratories.

## Data Availability

All data produced in the present study are available upon reasonable request to the authors

## List of abbreviations

CNS: *central nervous system*
CSF: *Cerebrospinal fluid*
FLCκ: *Free light chains kappa*
GFR: *glomerular filtration rate*
IF: *intrathecal fraction*
Ig: *Immunoglobulin*
OCB: *Oligoclonal bands*
QAlb: *Albumin quotient*
Qlim: *upper limit of the reference range*

## Declarations

Ethics approval and consent to participate

This study evaluates the sensitivity and specificity of a new laboratory parameter by using samples that would otherwise be discarded. The results are pseudonymized and not reported to the treating physician or the patient. Therefore no informed consent has to be obtained.

An ethics vote exists for this study, which approves the anonymized use of residual material for study purposes without informing the patients (BB135/22; ethics committee Universitymedicine Greifswald).

## Consent for publication

Not applicable

## Availability of data and materials

The datasets used and/or analysed during the current study are available from the corresponding author on reasonable request.

## Authors’ contributions

MS and AD have written the manuscript. All authors have contributed to the study design. All authors read and approved the final manuscript.

## Acknowledgements

Not applicable

